# Association between Albumin and Hearing Loss: A Cross-Sectional Study Using Data from the National Health and Nutrition Examination Survey (1999–2012, 2015–2018)

**DOI:** 10.1101/2025.08.13.25333551

**Authors:** Yiyang Chen, Chenyang Lei, Hongyi Lu, Wei Lin, Juan Jiang

## Abstract

**Objective:** We investigated the association between albumin levels and hearing loss.

**Methods:** A cross-sectional analysis was conducted using data from the National Health and Nutrition Examination Survey (NHANES) for the periods 1999–2012 and 2015–2018. Participants aged ≥20 years were included. Data on albumin levels, hearing ability, and relevant covariates were collected.

**Results:** The final sample comprised 12,133 adults with a mean age of 48.9 ± 17.5 years; 60.9% were non-Hispanic white, and 51.1% were men. Hearing loss was defined as a threshold of ≥20 dB. The odds ratios for low-frequency, speech-frequency, and high-frequency hearing loss were 0.69 (95% CI: 0.56–0.84, p < 0.001), 0.59 (95% CI: 0.49–0.73, p < 0.001), and 0.62 (95% CI: 0.51–0.76, p < 0.001), respectively.

**Conclusions:** These findings suggest that albumin levels are significantly associated with hearing loss in adults in the United States. However, further research into the underlying mechanisms is needed.

## Introduction

According to the World Health Organization (WHO), approximately 466 million individuals (6.1% of the global population) experienced disabling hearing loss (HL) in 2018[1]. Projections indicate that this number could exceed 700 million by 2030. Hearing loss is the fifth leading cause of years lived with disability[2]. Individuals with hearing loss are more likely to experience difficulties in daily activities, a reduced quality of life, and an increased risk of cognitive decline and depression[3–6]. The impact of hearing loss extends beyond individuals, affecting education, employment, socialization, and mental health while also placing a considerable burden on healthcare systems[7]. Because hearing loss is often irreversible, identifying potentially modifiable risk factors is a crucial public health objective. The etiology and pathophysiological mechanisms of hearing loss remain unclear and may involve microcirculatory disorders, viral infections, and autoimmune diseases[8]. Recent studies have also highlighted a strong association between hearing loss and cardiovascular disease[9,10]. Biological changes in liver function can reflect immune status and the ability of the body to repair damage. Serum albumin levels serve as indicators of nutritional status, with lower levels potentially leading to reduced immunity[11]. Serum albumin exhibits physiological properties, including anti-inflammatory, antioxidant, anticoagulant, and antiplatelet aggregation activities, as well as colloid osmotic effects [12,13]. Albumin interacts with vascular endothelium in multiple ways and may contribute to neuroprotection[14]. An increasing body of research supports the role of albumin in the development and progression of cardiovascular diseases[15,16]. Lower albumin levels within the normal range have been linked to the onset of sudden sensorineural hearing loss (SSNHL) and poor prognostic outcomes[17]. However, no study has specifically investigated the relationship between serum albumin levels and hearing loss in the general population.

Therefore, in this study, we aimed to examine the clinical and prognostic relevance of albumin in individuals with hearing loss within a specified study cohort. A retrospective cross-sectional study was conducted, including 12,133 participants from the National Health and Nutrition Examination Survey (NHANES).

## Materials and Methods

### Study Population

This cross-sectional study used data from the National Health and Nutrition Examination Survey (NHANES) spanning 1999–2012 and 2015–2018, conducted by the Centers for Disease Control and Prevention (CDC)[18]. NHANES evaluates the health and nutritional status of non-hospitalized Americans. Data collection involved demographic surveys, comprehensive health assessments conducted during home visits, and laboratory tests performed at mobile screening centers. The National Center for Health Statistics Ethics Review Committee approved the NHANES protocol, and all participants provided written informed consent before participation. Because this study was a secondary analysis, additional Institutional Review Board approval was not required[19]. The NHANES data used in this study were accessed on March 1, 2022, via the CDC website (http://www.cdc.gov/nchs/nhanes.htm). This study included individuals aged 20 years or older who completed the interviews. Participants with missing audiometric data, albumin levels, or key covariates were excluded. The total sample size was 49,312 adults, as shown in Figure 1. Only publicly available data were used; thus, ethical approval was not required.

**Figure 1.**
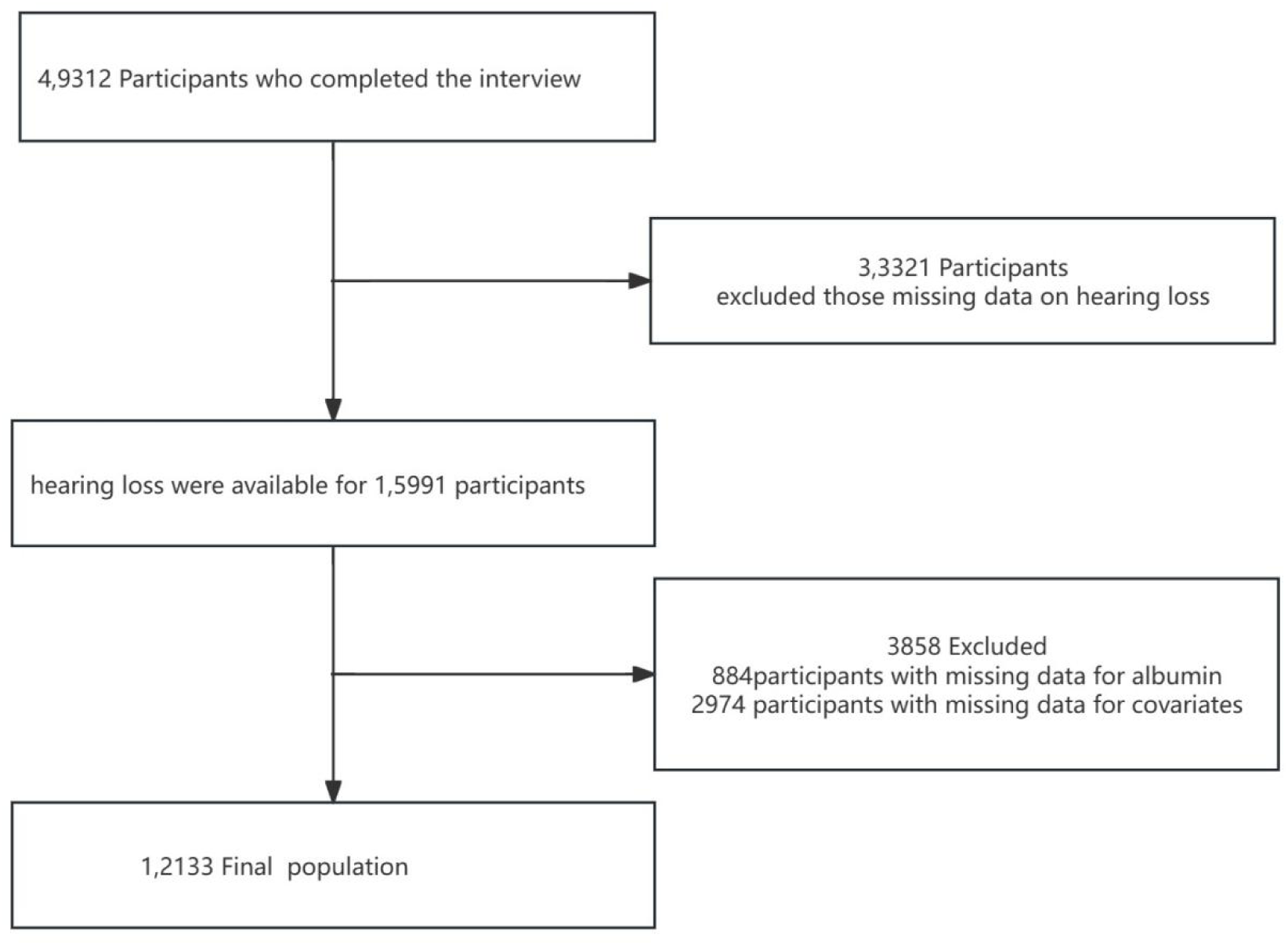
the study’s flow diagram

### Audiometric Measurements and Definition of Hearing Loss

Hearing tests for participants aged ≥20 years were conducted by trained examiners in a dedicated soundproof room at a mobile testing center. Binaural hearing thresholds were measured at seven frequencies (500, 1000, 2000, 3000, 4000, 6000, and 8000 Hz), with results ranging from −10 to 120 dB. Participants with abnormal otoscopy or tympanogram results were excluded. The pure tone average (PTA) for speech frequencies was calculated as the average of hearing thresholds at 0.5, 1, 2, and 4 kHz. The better ear PTA has been reported as a continuous variable, with higher values indicating poorer hearing. The low-frequency PTA was calculated using thresholds of 0.5, 1, and 2 kHz, whereas the high-frequency PTA was calculated using thresholds of 4, 6, and 8 kHz. All hearing thresholds are expressed in dB HL. Sensitivity analyses of PTA were conducted using the worse ear rather than the better ear. According to the World Health Organization’s 2021 definition, hearing loss is classified as a speech frequency PTA of ≥20 dB in the better ear, whereas <20 dB is considered normal hearing[20,21].

### Measurement of Albumin

Laboratory personnel, including medical technicians and phlebotomists, underwent comprehensive training in standardized laboratory procedures before commencing work at the Mobile Examination Center (MEC). Albumin concentration was measured using the bromocresol purple (BCP) dye-binding method. A color change at 600 nm occurred when the dye selectively bound to albumin within a pH range of 5.2–6.8. A secondary wavelength of 700 nm, which specifically targets albumin, was used for the endpoint reaction.

### Covariates

Multiple covariates were assessed based on prior literature[20,22,23], including age, sex, race, marital status, household income, education level, smoking status, alcohol use, physical activity, body mass index (BMI), hypertension, diabetes, noise exposure, alanine aminotransferase, glutamine aminotransferase, and total protein levels. Race/ethnicity was categorized as Mexican American, other Hispanic, non-Hispanic white, non-Hispanic black, or other (including multiple races). Marital status was categorized as married, unmarried, cohabiting, widowed, divorced, or separated. Education level was classified as less than 9th grade, 9th– 11th grade (including 12th grade without a diploma), high school graduate/general educational development (GED) or equivalent, college or Associate of Arts (AA) degree, and college graduate or higher.

Smoking status was categorized as never smoked (fewer than 100 cigarettes), current smoker, or former smoker (quit smoking after smoking more than 100 cigarettes). Physical activity was classified as sedentary, moderate (≥10 minutes of exercise in the past 30 days leading to light sweating or a mild-to-moderate increase in respiration or heart rate), or vigorous (≥10 minutes of exercise in the past 30 days resulting in heavy sweating or a substantial increase in respiration or heart rate). Hypertension and diabetes were determined based on self-reported physician diagnoses. BMI was calculated using standardized techniques based on weight and height. Noise exposure assessments included nonworkplace noise, gunfire exposure outside the workplace, workplace noise, and working in noisy environments.

This study involved a secondary analysis of publicly accessible datasets, with categorical variables expressed as proportions (%) and continuous variables expressed as means (standard deviation [SD]) or medians (interquartile range [IQR]). One-way analysis of variance (ANOVA) was used for normally distributed variables, chi-square tests for categorical variables, and Kruskal-Wallis tests for skewed distributions. Logistic regression models were employed to calculate odds ratios (ORs) and 95% confidence intervals (CIs) to assess the association between albumin levels and hearing loss, adjusting for all covariates. Model 1 was unadjusted. Model 2 adjusted for sociodemographic characteristics, including age, sex, race, marital status, household income, education, smoking status, alcohol consumption, and physical activity. Model 3 additionally adjusted for BMI, hypertension, diabetes, noise exposure, alanine aminotransferase, glutamine aminotransferase, and total protein levels.

All analyses were conducted using R software version 4.3.1 (http://www.R-project.org; R Foundation, Shanghai, China) (accessed on March 10, 2024) and Free Statistics software version 1.9. Descriptive statistics were calculated for all participants, and statistical significance was set at *p* < 0.05 for two-tailed tests.

## Results

### Study Population

A total of 49,312 participants aged 20 years or older completed the interviews. Participants with missing data on hearing loss (*n* = 33,321), albumin levels (*n* = 884), or other covariates (*n* = 2,974) were excluded. Ultimately, 12,133 participants from the NHANES conducted between 1999–2012 and 2015–2018 were included in this cross-sectional study. The detailed inclusion and exclusion criteria are shown in Figure 1.

### Baseline Characteristics

Table 1 summarizes the baseline characteristics of participants categorized by albumin tertiles. The mean age of participants was 48.9 (±17.5) years, and 5,929 (48.9%) were women. Participants with higher albumin levels were more likely to be non-Hispanic white, married, have some college education or an associate degree, never smoke, consume alcohol in moderation, and have a lower prevalence of diabetes and hypertension. They also had lower rates of low-frequency, speech-frequency, and high-frequency hearing loss, as well as reduced noise exposure.

**Table 1.**
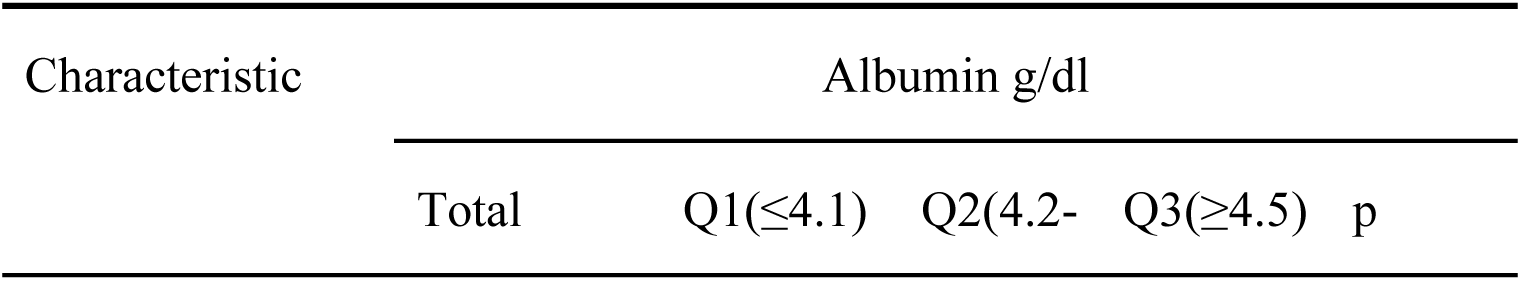
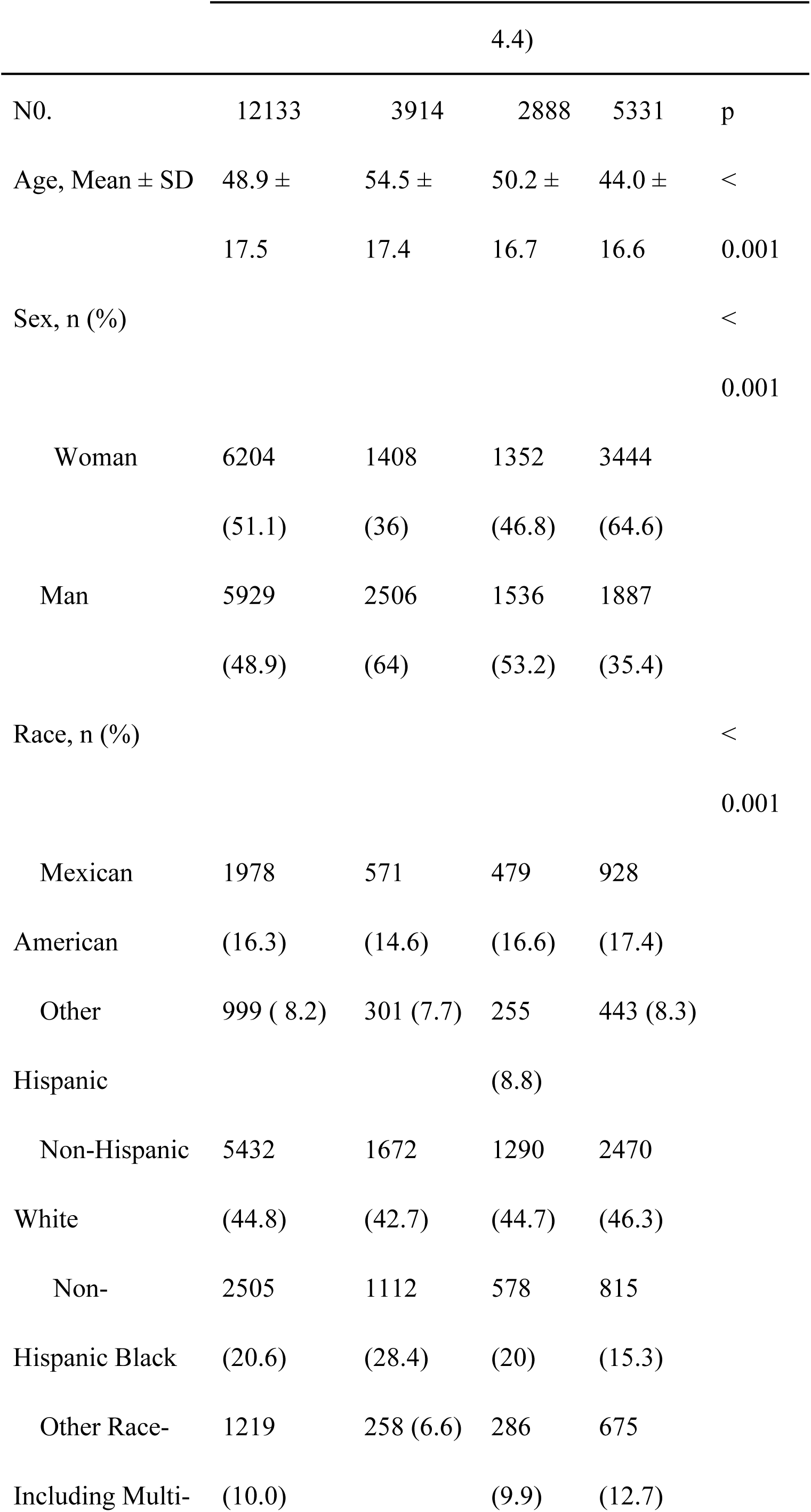
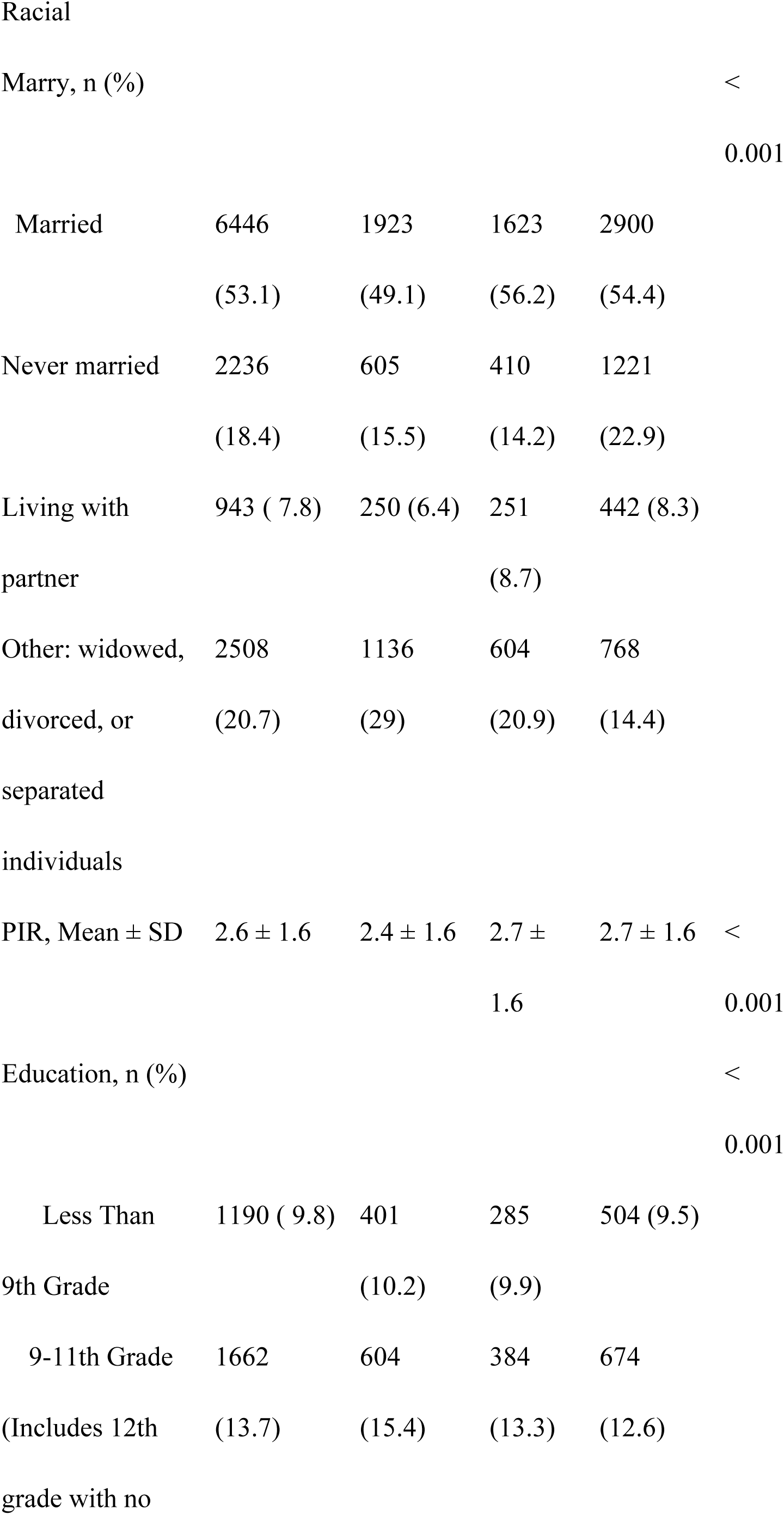
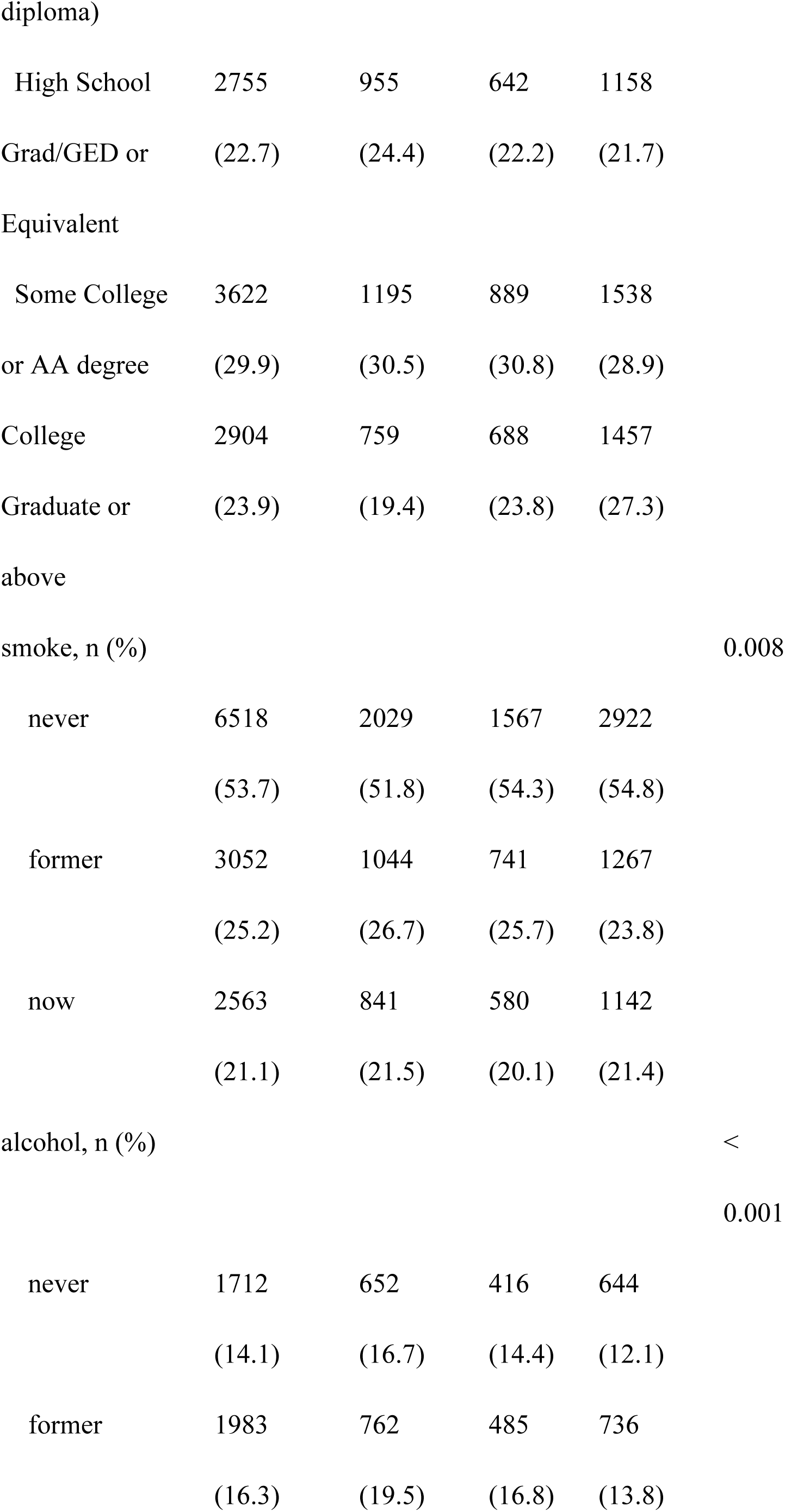
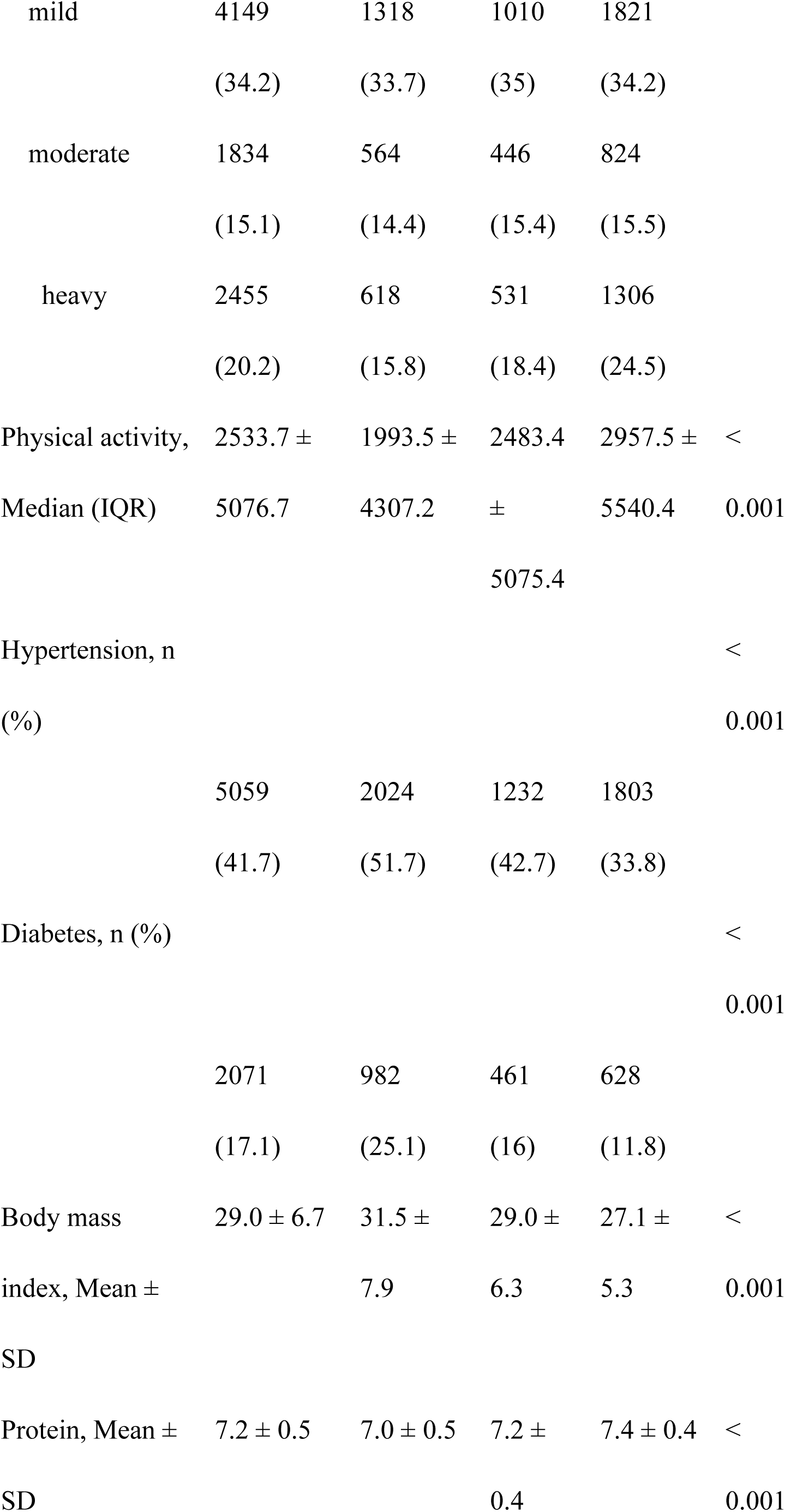
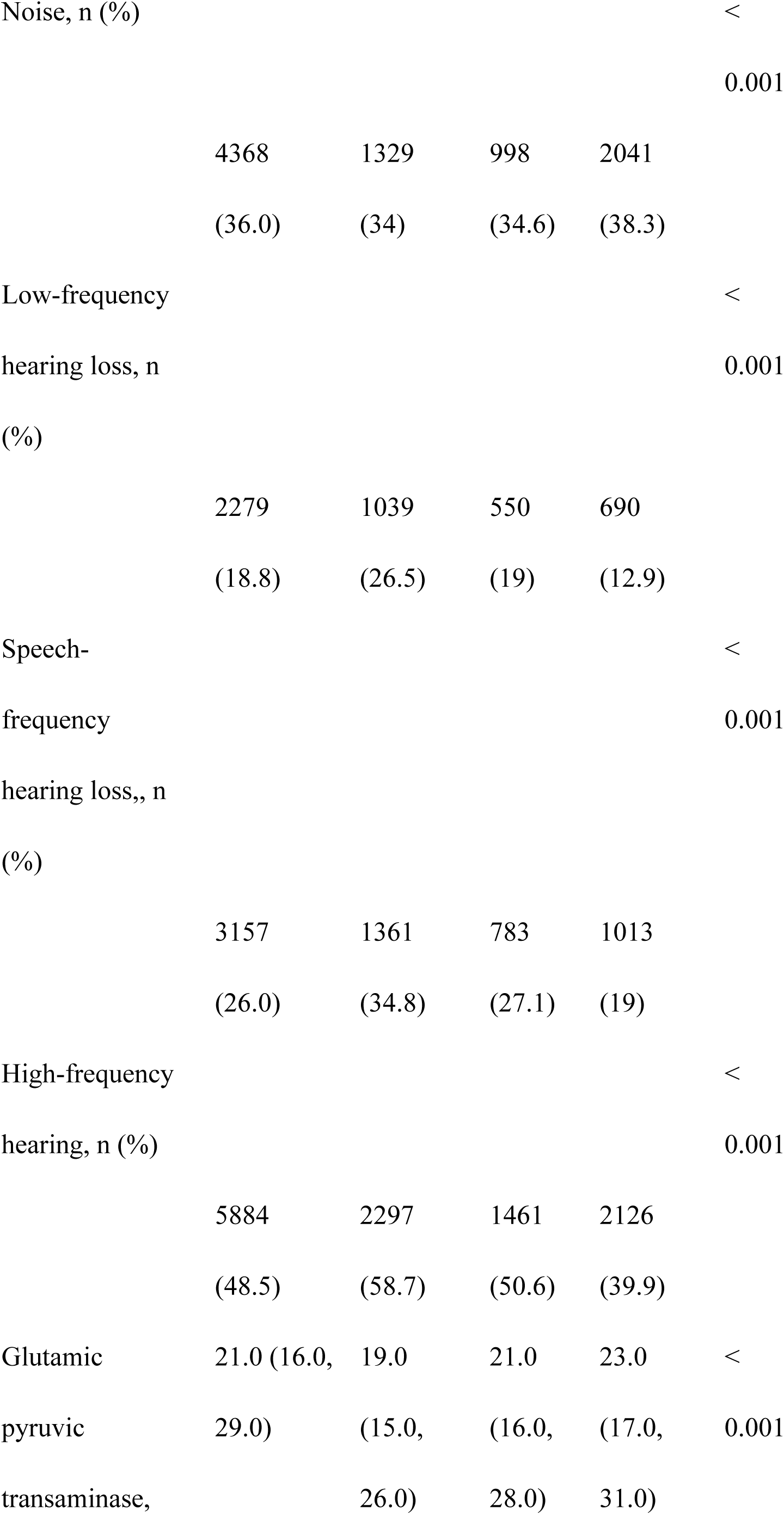
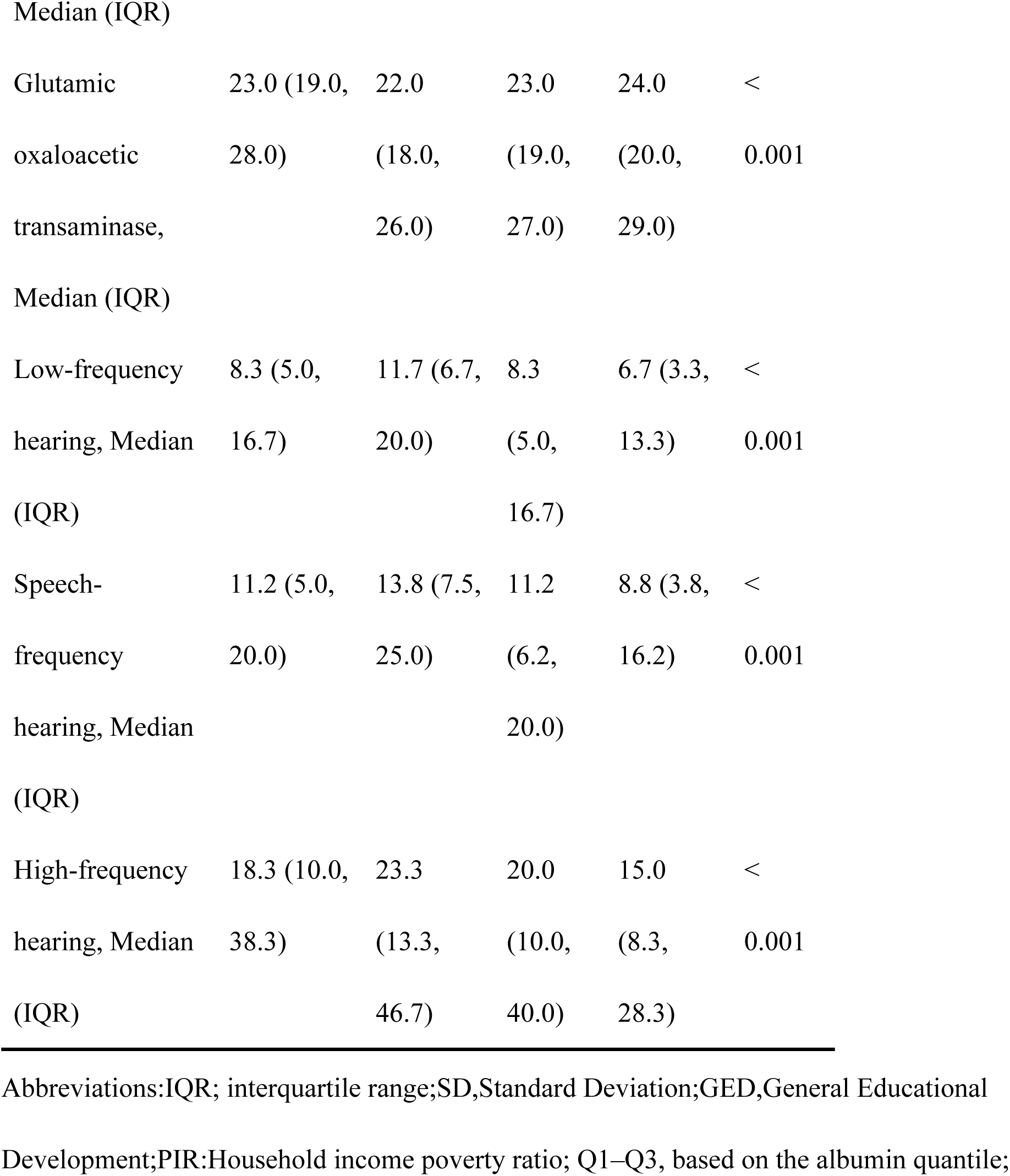
Population characteristics by categories of albumin.

### Relationship between Albumin and Hearing Loss

Univariate analyses indicated associations between hearing loss and age, sex, race, marital status, household income, education, smoking status, alcohol consumption, physical activity, hypertension, diabetes, noise exposure, and albumin levels (Table 2). When albumin levels were analyzed by tertiles, a significant negative correlation was observed between plasma albumin levels and hearing loss, even after adjusting for potential confounders(Table 3). For low-frequency hearing loss, the adjusted odds ratios (ORs) for albumin levels in Q2 (4.2–4.4 g/dl) and Q3 (≥4.5 g/dl) compared with Q1 (≤4.0 g/dl) were 0.88 (95% CI: 0.76–1.02, *p* = 0.086) and 0.79 (95% CI: 0.68–0.92, *p* = 0.002), respectively. For speech-frequency hearing loss, the adjusted ORs were 0.88 (95% CI: 0.76–1.01, *p* = 0.076) and 0.73 (95% CI: 0.63–0.85, *p* < 0.001). For high-frequency hearing loss, the adjusted ORs were 0.81 (95% CI: 0.70–0.94, *p* = 0.04) and 0.72 (95% CI: 0.62–0.83, *p* < 0.001), respectively. When albumin was analyzed as a continuous variable, each 1 g/dl increase in plasma albumin was associated with a 21% reduction in the risk of low-frequency hearing loss (OR: 0.69, 95% CI: 0.56–0.84, *p* < 0.001), a 31% reduction in the risk of speech-frequency hearing loss (OR: 0.59, 95% CI: 0.49–0.73, *p* < 0.001), and a 38% reduction in the risk of high-frequency hearing loss (OR: 0.62, 95% CI: 0.51–0.76, *p* < 0.001).

**Table 2.**
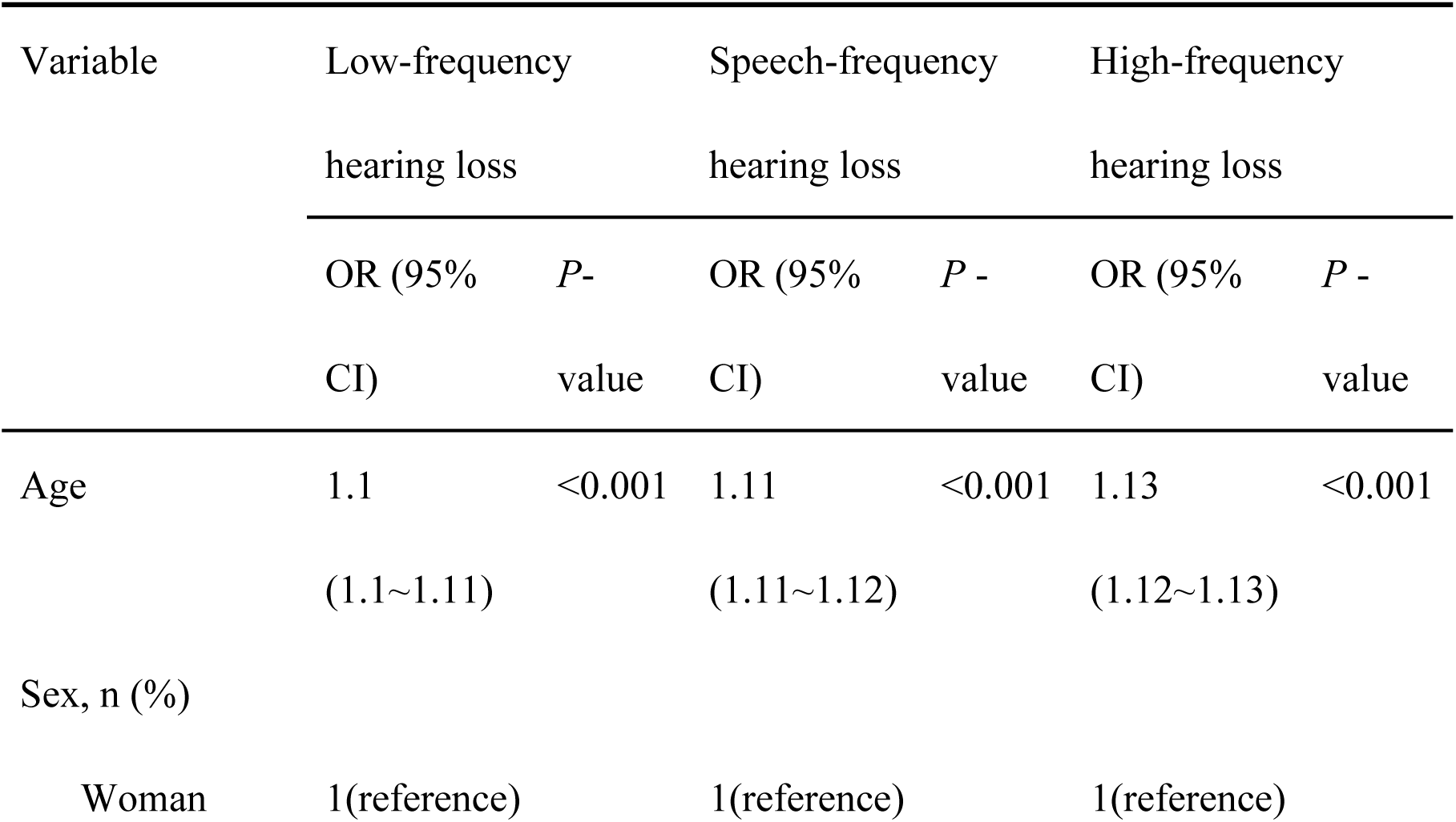
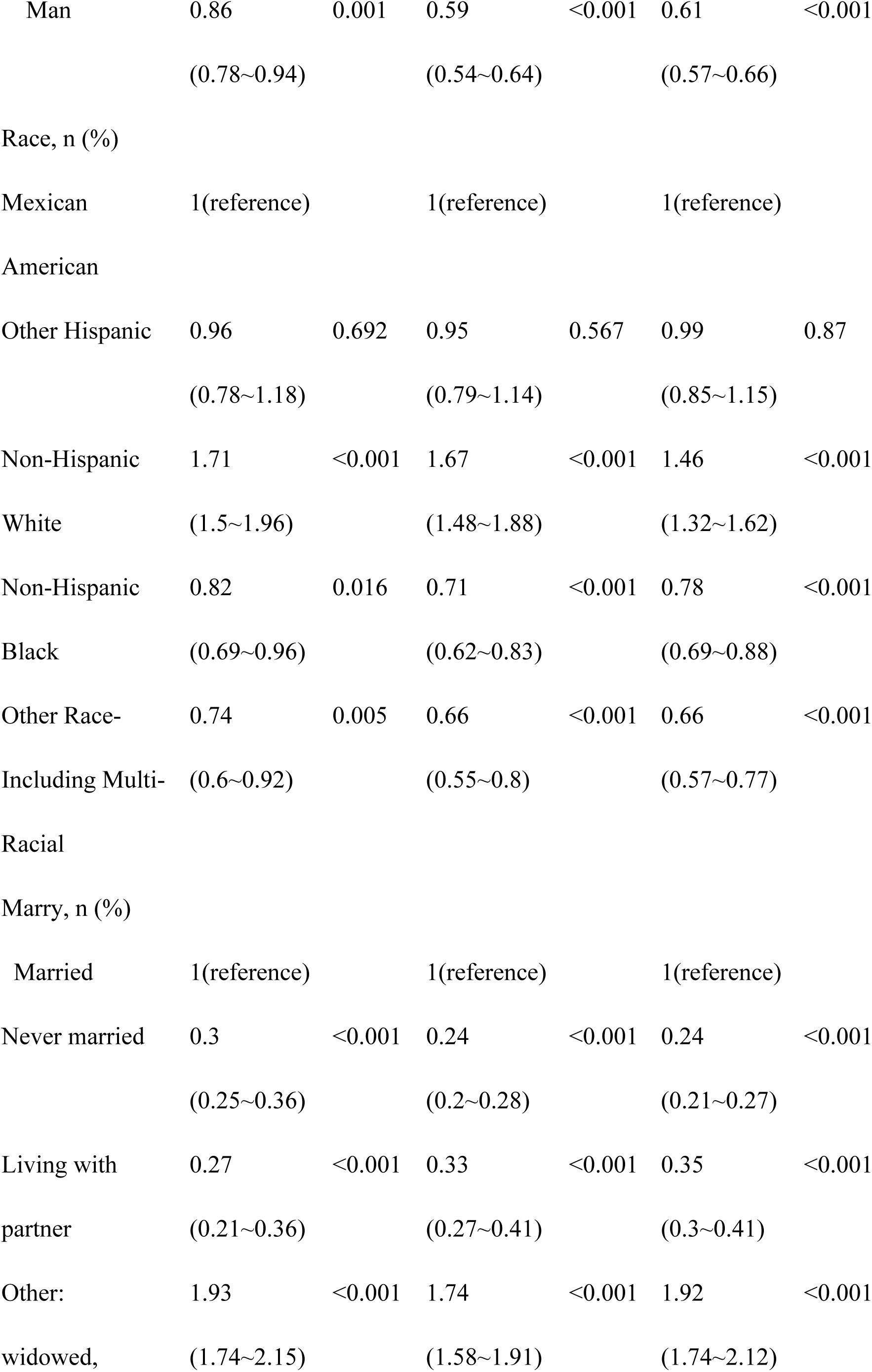
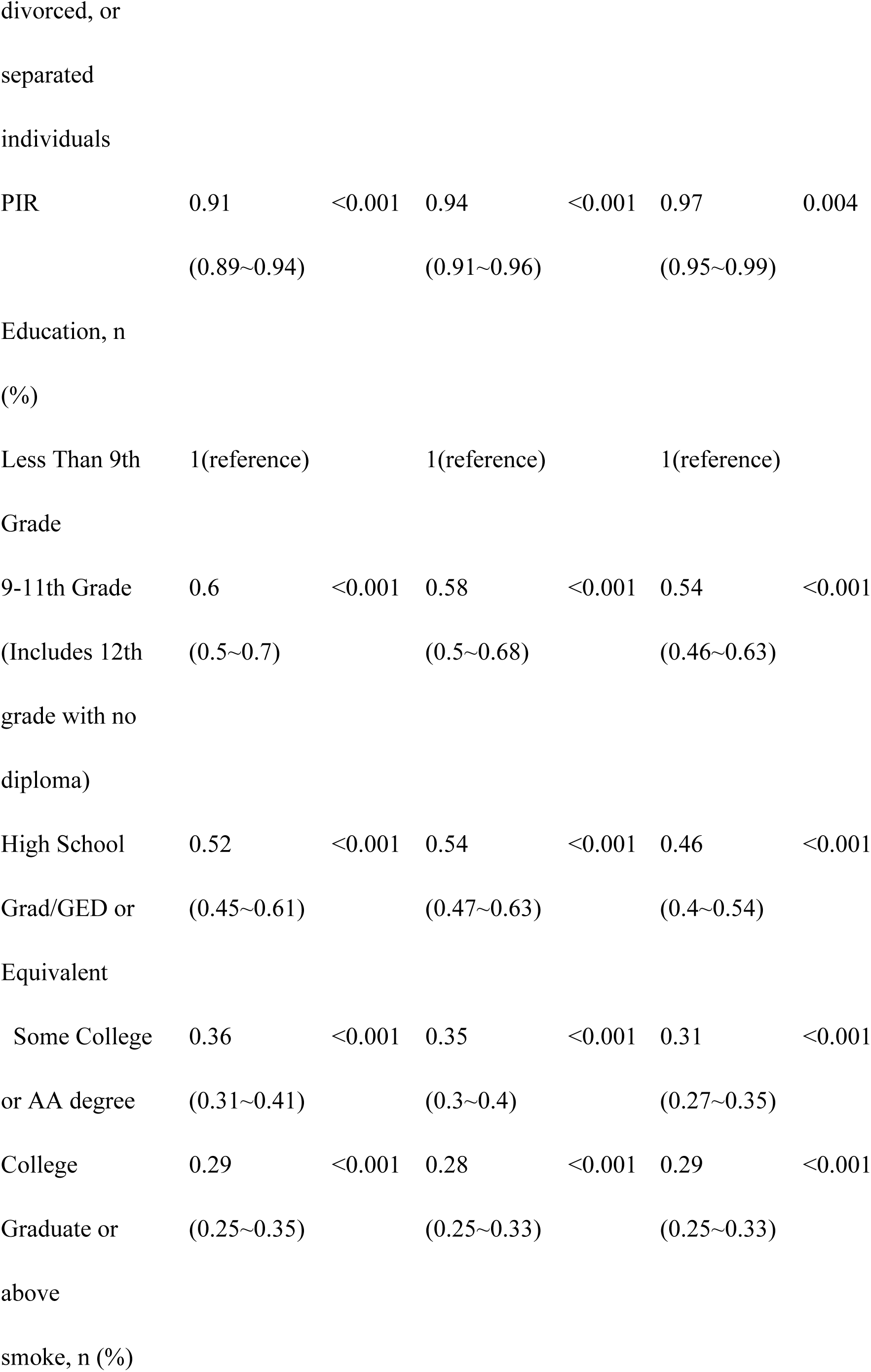
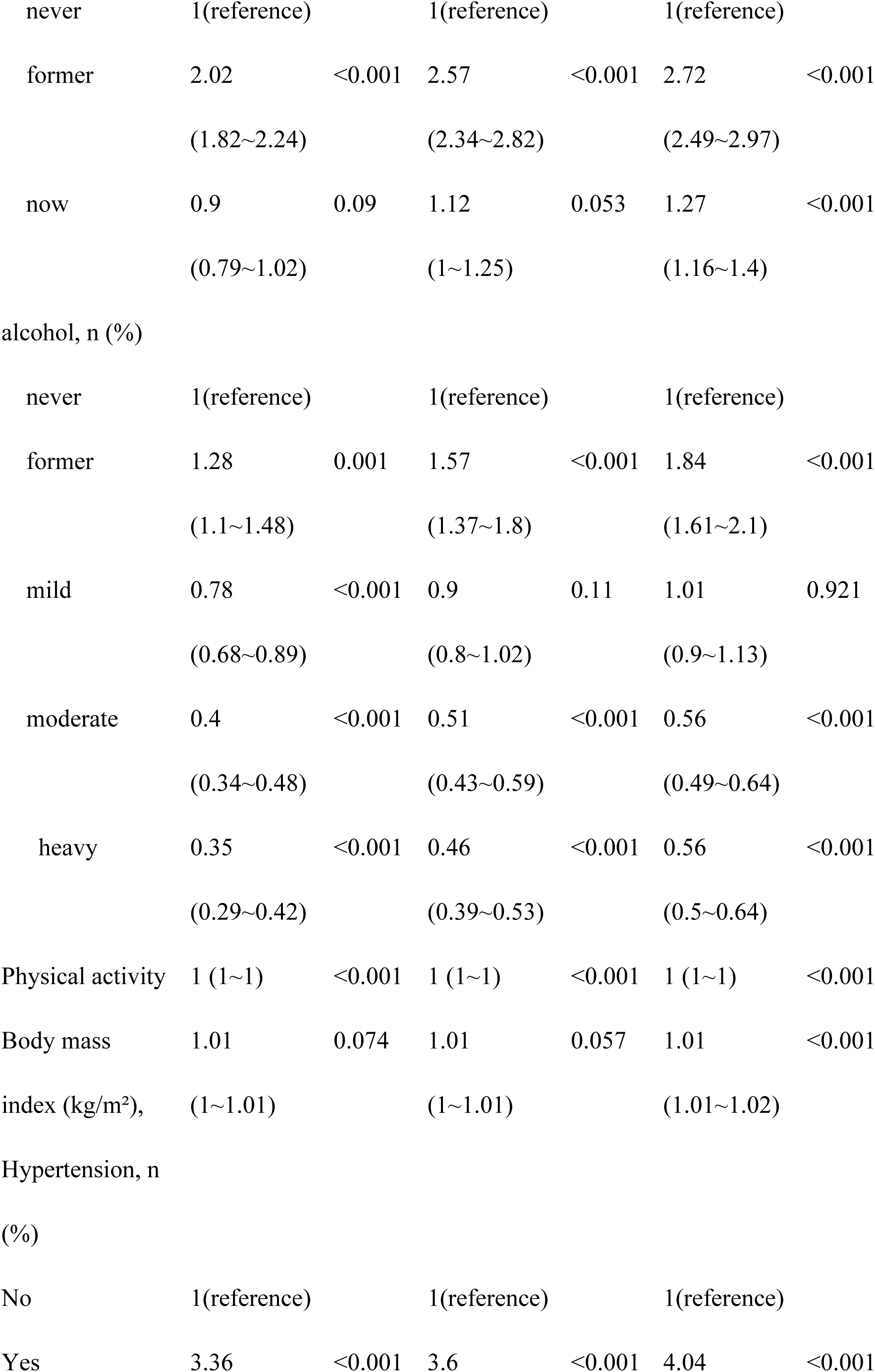
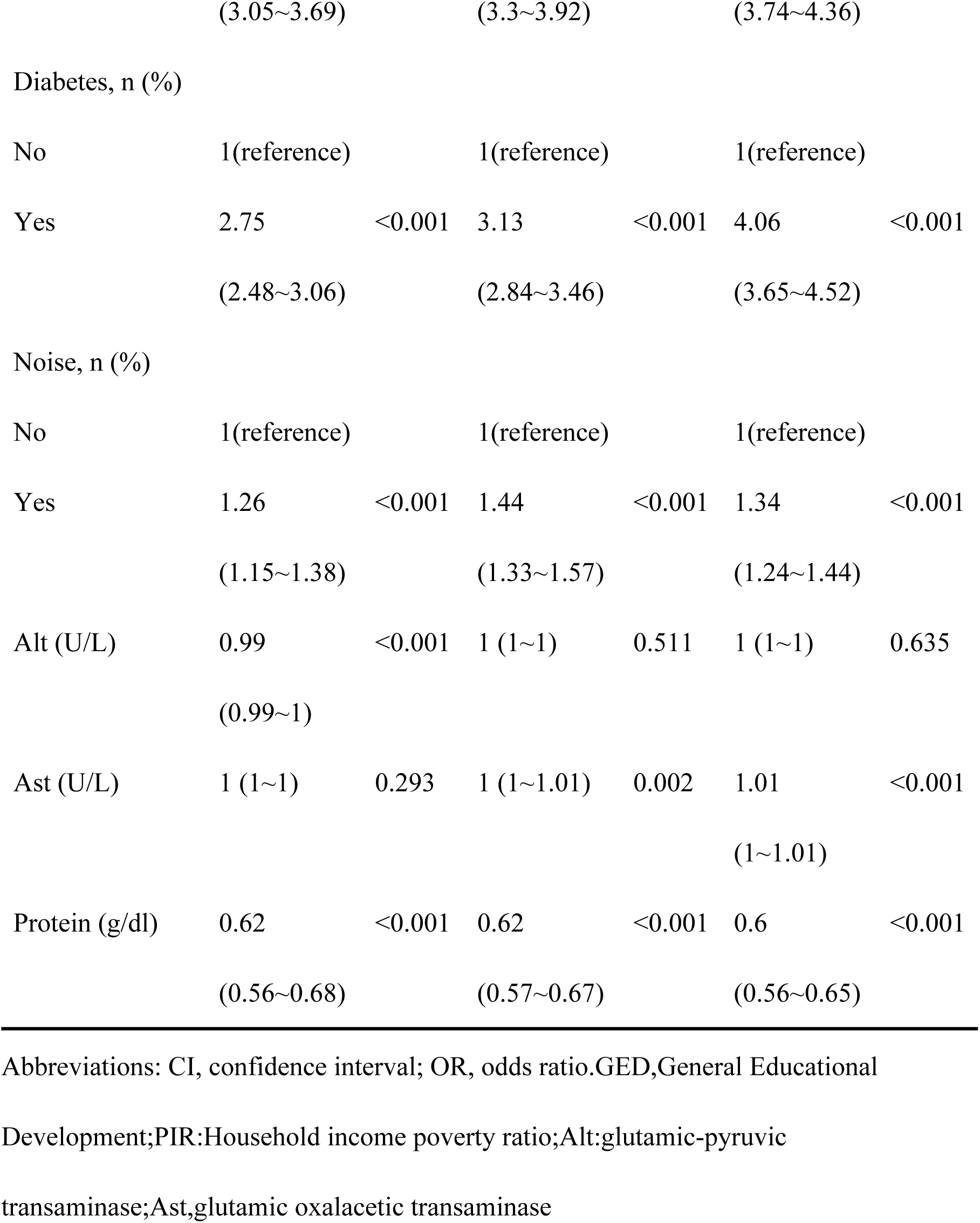
Association of covariates and hearing loss.

| Variable | Low-frequency |  | Speech-frequency |  | High-frequency |  |
| --- | --- | --- | --- | --- | --- | --- |
|  | hearing loss |  | hearing loss |  | hearing loss |  |
|  | OR (95% CI) | *P*-value | OR (95% CI) | *P*-value | OR (95% CI) | *P*-value |
| Age | 1.1<br>(1.1~1.11) | <0.001 | 1.11<br>(1.11~1.12) | <0.001 | 1.13<br>(1.12~1.13) | <0.001 |
| Sex, n (%) |  |  |  |  |  |  |
| Woman | 1(reference) |  | 1(reference) |  | 1(reference) |  |
| Man | 0.86 | 0.001 | 0.59 | <0.001 | 0.61 | <0.001 |
|  | (0.78~0.94) |  | (0.54~0.64) |  | (0.57~0.66) |  |
| Race, n (%) |  |  |  |  |  |  |
| Mexican | 1(reference) |  | 1(reference) |  | 1(reference) |  |
| American |  |  |  |  |  |  |
| Other Hispanic | 0.96 | 0.692 | 0.95 | 0.567 | 0.99 | 0.87 |
|  | (0.78~1.18) |  | (0.79~1.14) |  | (0.85~1.15) |  |
| Non-Hispanic | 1.71 | <0.001 | 1.67 | <0.001 | 1.46 | <0.001 |
| White | (1.5~1.96) |  | (1.48~1.88) |  | (1.32~1.62) |  |
| Non-Hispanic | 0.82 | 0.016 | 0.71 | <0.001 | 0.78 | <0.001 |
| Black | (0.69~0.96) |  | (0.62~0.83) |  | (0.69~0.88) |  |
| Other Race- | 0.74 | 0.005 | 0.66 | <0.001 | 0.66 | <0.001 |
| Including Multi- | (0.6~0.92) |  | (0.55~0.8) |  | (0.57~0.77) |  |
| Racial |  |  |  |  |  |  |
| Marry, n (%) |  |  |  |  |  |  |
| Married | 1(reference) |  | 1(reference) |  | 1(reference) |  |
| Never married | 0.3 | <0.001 | 0.24 | <0.001 | 0.24 | <0.001 |
|  | (0.25~0.36) |  | (0.2~0.28) |  | (0.21~0.27) |  |
| Living with | 0.27 | <0.001 | 0.33 | <0.001 | 0.35 | <0.001 |
| partner | (0.21~0.36) |  | (0.27~0.41) |  | (0.3~0.41) |  |
| Other: | 1.93 | <0.001 | 1.74 | <0.001 | 1.92 | <0.001 |
| widowed, | (1.74~2.15) |  | (1.58~1.91) |  | (1.74~2.12) |  |
| PIR | 0.91 | <0.001 | 0.94 | <0.001 | 0.97 | 0.004 |
|  | (0.89~0.94) |  | (0.91~0.96) |  | (0.95~0.99) |  |

|  |  |  |  |  |  |
| --- | --- | --- | --- | --- | --- |
| Less Than 9th | 1(reference) |  | 1(reference) |  | 1(reference) |
| Grade |  |  |  |  |  |  |
| 9-11th Grade | 0.6 | <0.001 | 0.58 | <0.001 | 0.54 | <0.001 |
| (Includes 12th | (0.5~0.7) |  | (0.5~0.68) |  | (0.46~0.63) |  |
| High School | 0.52 | <0.001 | 0.54 | <0.001 | 0.46 | <0.001 |
| Grad/GED or | (0.45~0.61) |  | (0.47~0.63) |  | (0.4~0.54) |  |
| Equivalent |  |  |  |  |  |  |
| Some College | 0.36 | <0.001 | 0.35 | <0.001 | 0.31 | <0.001 |
| or AA degree | (0.31~0.41) |  | (0.3~0.4) |  | (0.27~0.35) |  |
| College | 0.29 | <0.001 | 0.28 | <0.001 | 0.29 | <0.001 |
| Graduate or | (0.25~0.35) |  | (0.25~0.33) |  | (0.25~0.33) |  |
| never | 1(reference) |  | 1(reference) |  | 1(reference) |  |
| former | 2.02 | <0.001 | 2.57 | <0.001 | 2.72 | <0.001 |
|  | (1.82~2.24) |  | (2.34~2.82) |  | (2.49~2.97) |  |
| now | 0.9 | 0.09 | 1.12 | 0.053 | 1.27 | <0.001 |
|  | (0.79~1.02) |  | (1~1.25) |  | (1.16~1.4) |  |
| alcohol, n (%) |  |  |  |  |  |  |
| never | 1(reference) |  | 1(reference) |  | 1(reference) |  |
| former | 1.28 | 0.001 | 1.57 | <0.001 | 1.84 | <0.001 |
|  | (1.1~1.48) |  | (1.37~1.8) |  | (1.61~2.1) |  |
| mild | 0.78 | <0.001 | 0.9 | 0.11 | 1.01 | 0.921 |
|  | (0.68~0.89) |  | (0.8~1.02) |  | (0.9~1.13) |  |
| moderate | 0.4 | <0.001 | 0.51 | <0.001 | 0.56 | <0.001 |
|  | (0.34~0.48) |  | (0.43~0.59) |  | (0.49~0.64) |  |
| heavy | 0.35 | <0.001 | 0.46 | <0.001 | 0.56 | <0.001 |
|  | (0.29~0.42) |  | (0.39~0.53) |  | (0.5~0.64) |  |
| Physical activity | 1 (1~1) | <0.001 | 1 (1~1) | <0.001 | 1 (1~1) | <0.001 |
| Body mass | 1.01 | 0.074 | 1.01 | 0.057 | 1.01 | <0.001 |
| index (kg/m<sup>2</sup>), | (1~1.01) |  | (1~1.01) |  | (1.01~1.02) |  |
| Hypertension, n (%) |  |  |  |  |  |  |
| No | 1(reference) |  | 1(reference) |  | 1(reference) |  |
| Yes | 3.36 | <0.001 | 3.6 | <0.001 | 4.04 | <0.001 |
|  | (3.05~3.69) |  | (3.3~3.92) |  | (3.74~4.36) |  |
| Diabetes, n (%) |  |  |  |  |  |  |
| No | 1(reference) |  | 1(reference) |  | 1(reference) |  |
| Yes | 2.75 | <0.001 | 3.13 | <0.001 | 4.06 | <0.001 |
|  | (2.48~3.06) |  | (2.84~3.46) |  | (3.65~4.52) |  |
| Noise, n (%) |  |  |  |  |  |  |
| No | 1(reference) |  | 1(reference) |  | 1(reference) |  |
| Yes | 1.26 | <0.001 | 1.44 | <0.001 | 1.34 | <0.001 |
|  | (1.15~1.38) |  | (1.33~1.57) |  | (1.24~1.44) |  |
| Alt (U/L) | 0.99 | <0.001 | 1 (1~1) | 0.511 | 1 (1~1) | 0.635 |
|  | (0.99~1) |  |  |  |  |  |
| Ast (U/L) | 1 (1~1) | 0.293 | 1 (1~1.01) | 0.002 | 1.01 | <0.001 |
|  |  |  |  |  | (1~1.01) |  |
| Protein (g/dl) | 0.62 | <0.001 | 0.62 | <0.001 | 0.6 | <0.001 |
|  | (0.56~0.68) |  | (0.57~0.67) |  | (0.56~0.65) |  |

Abbreviations: CI, confidence interval; OR, odds ratio.GED,General Educational
Development;PIR:Household income poverty ratio;Alt:glutamic-pyruvic
transaminase;Ast,glutamic oxalacetic transaminase

**Table 3.**
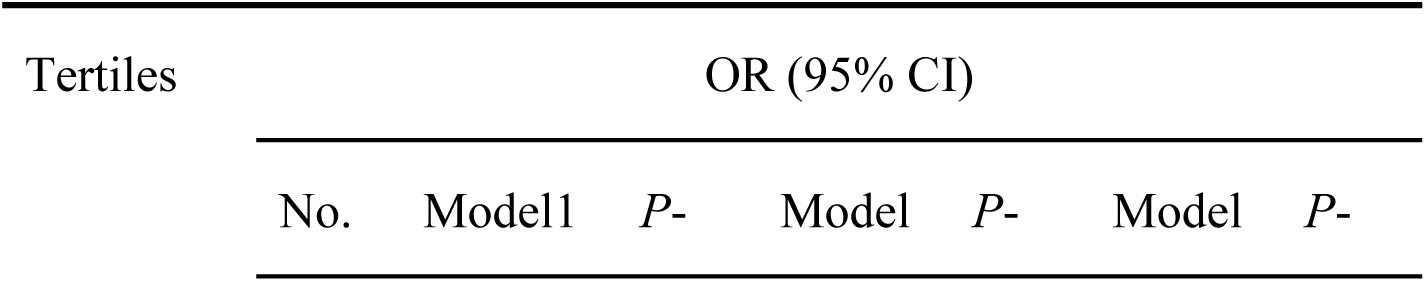
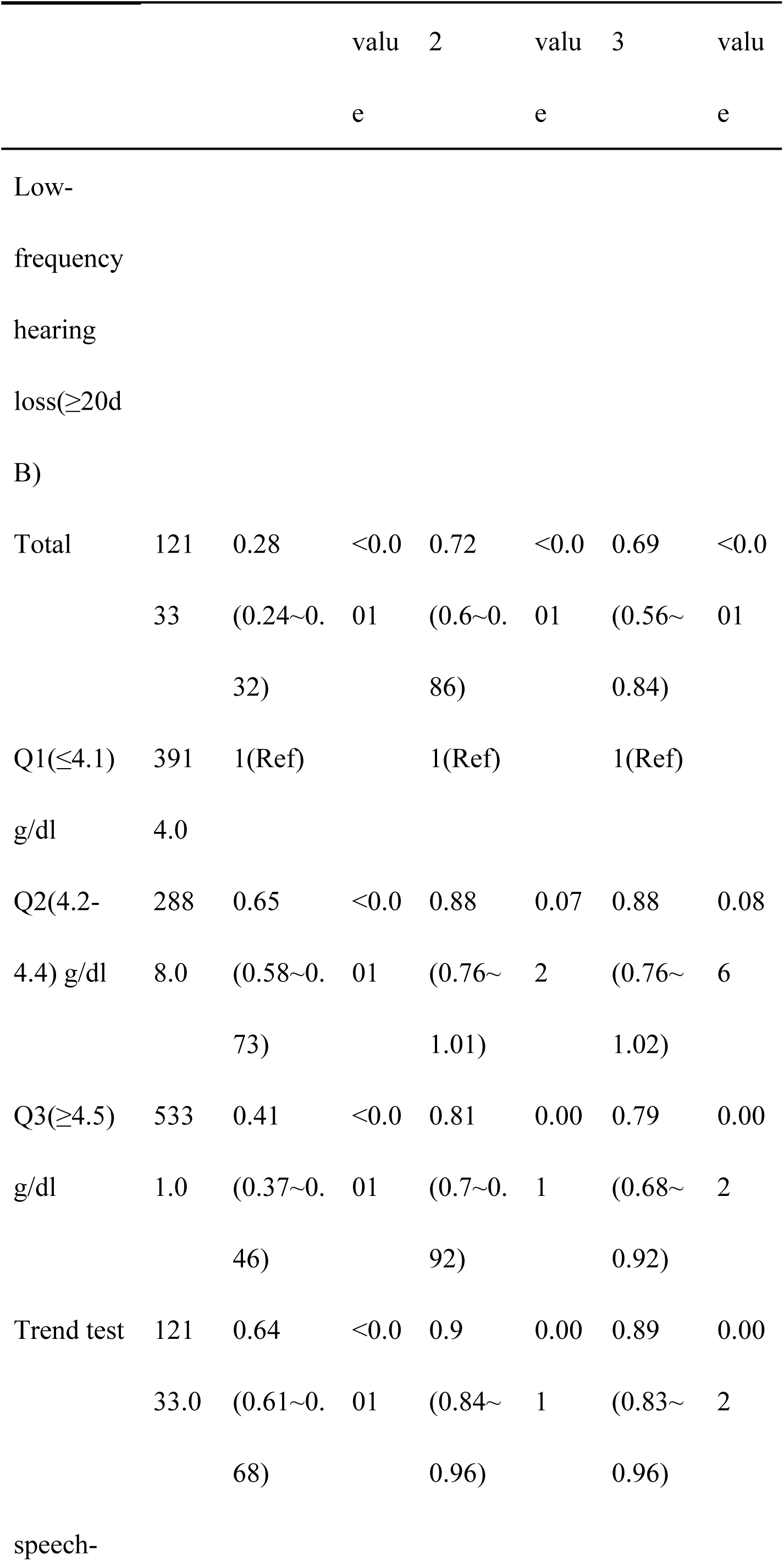
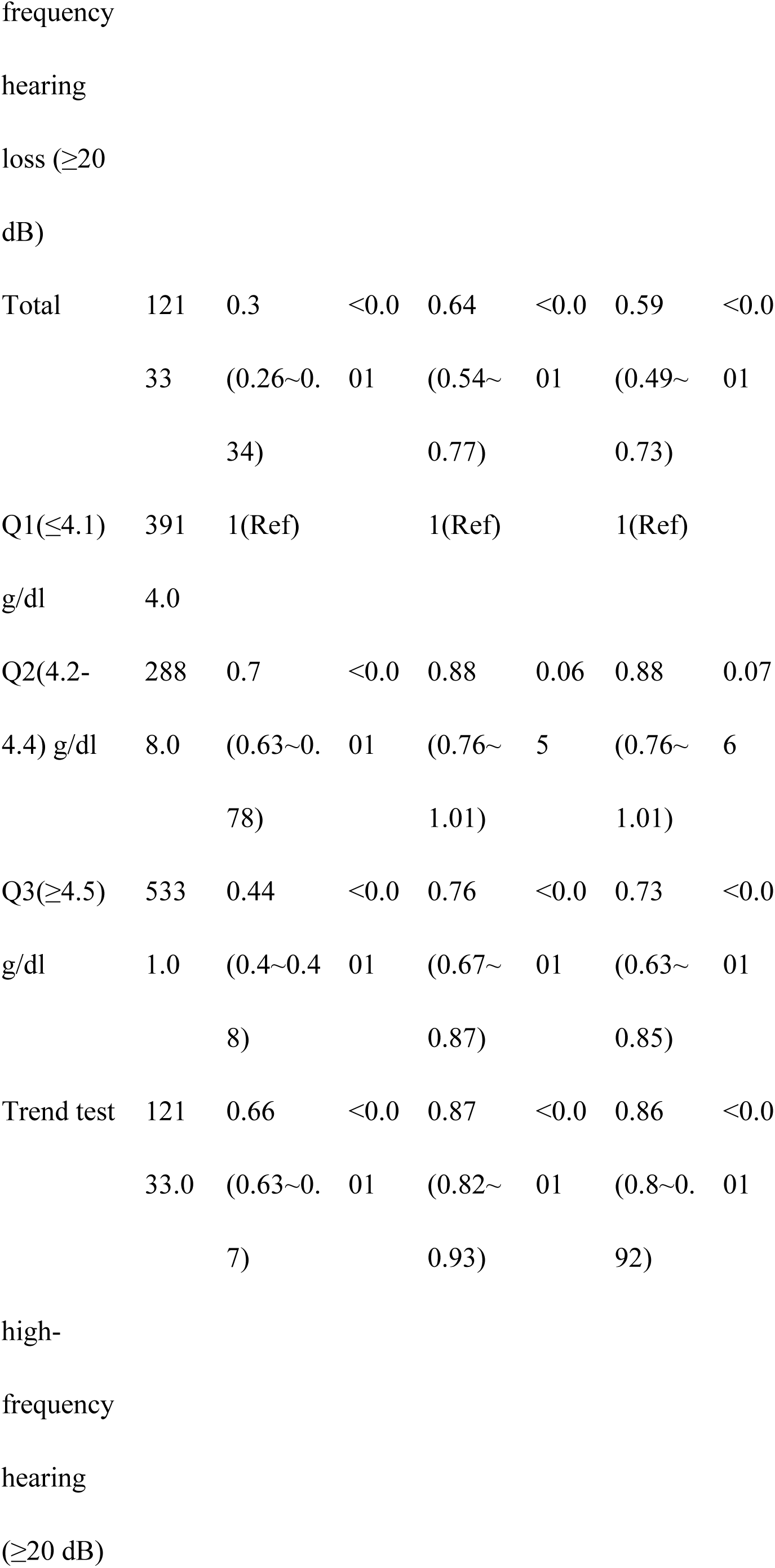
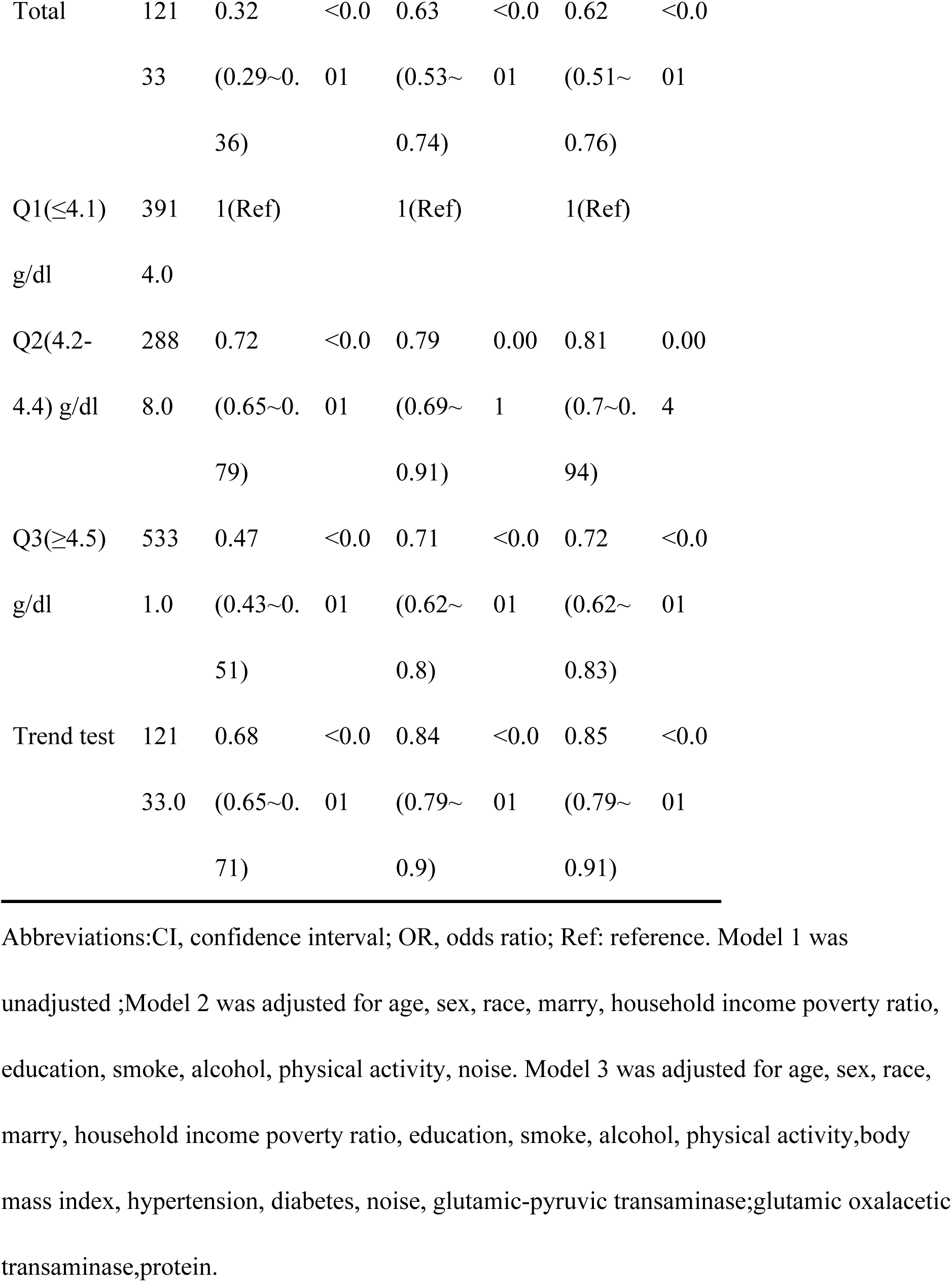
Association between albumin and hearing loss.

### Stratified Analyses Based on Additional Variables

Stratified analyses were conducted across various subgroups to assess the potential influence of plasma albumin levels on hearing loss. No significant interactions were observed after stratification by sex, age, BMI, household income, marital status, hypertension, or diabetes mellitus (Figure 2,3,4).

**Figure 2.**
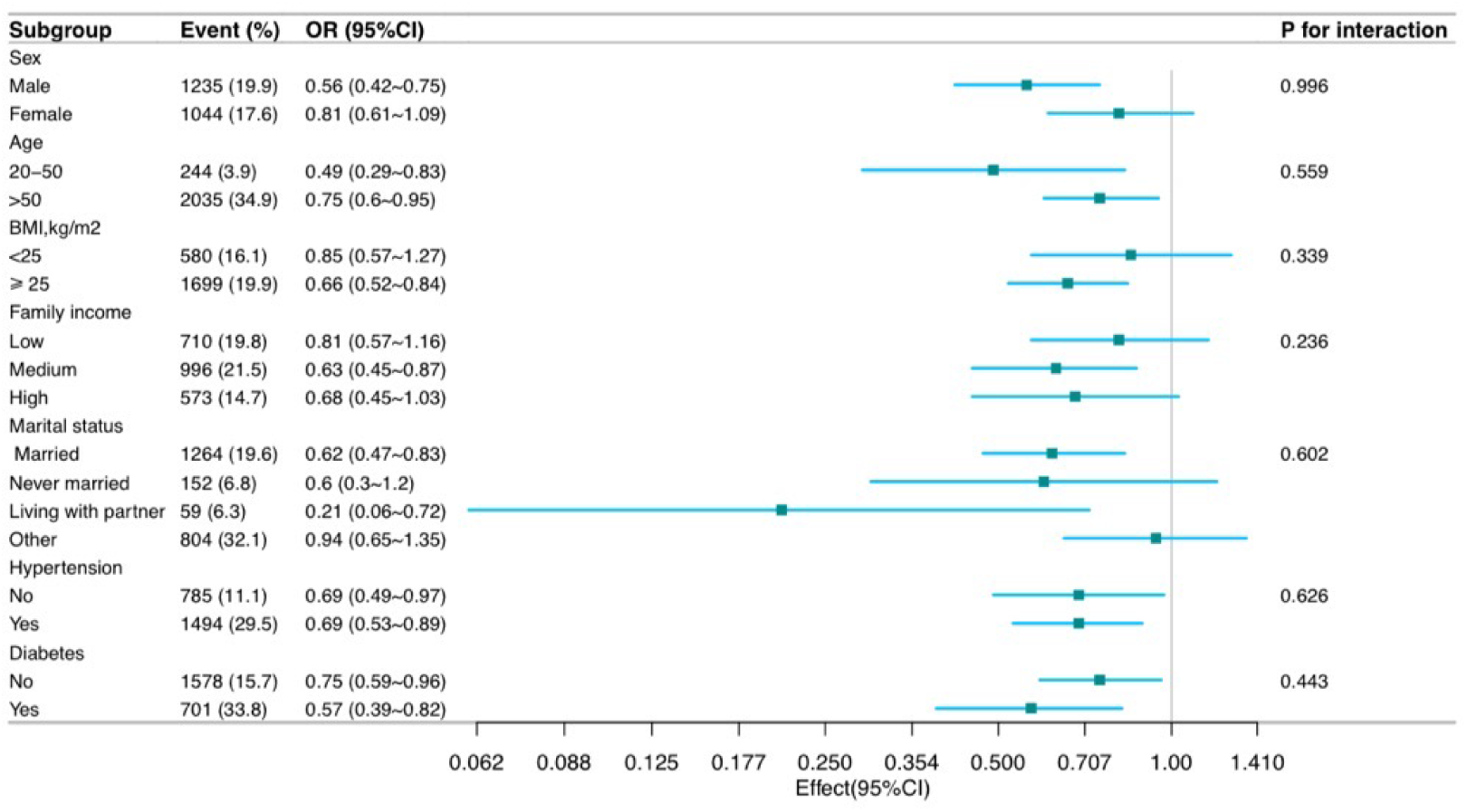
Relationship between albumin and Low-frequency hearing loss Abbreviations:CI, confidence interval; OR, odds ratio.

**Figure 3.**
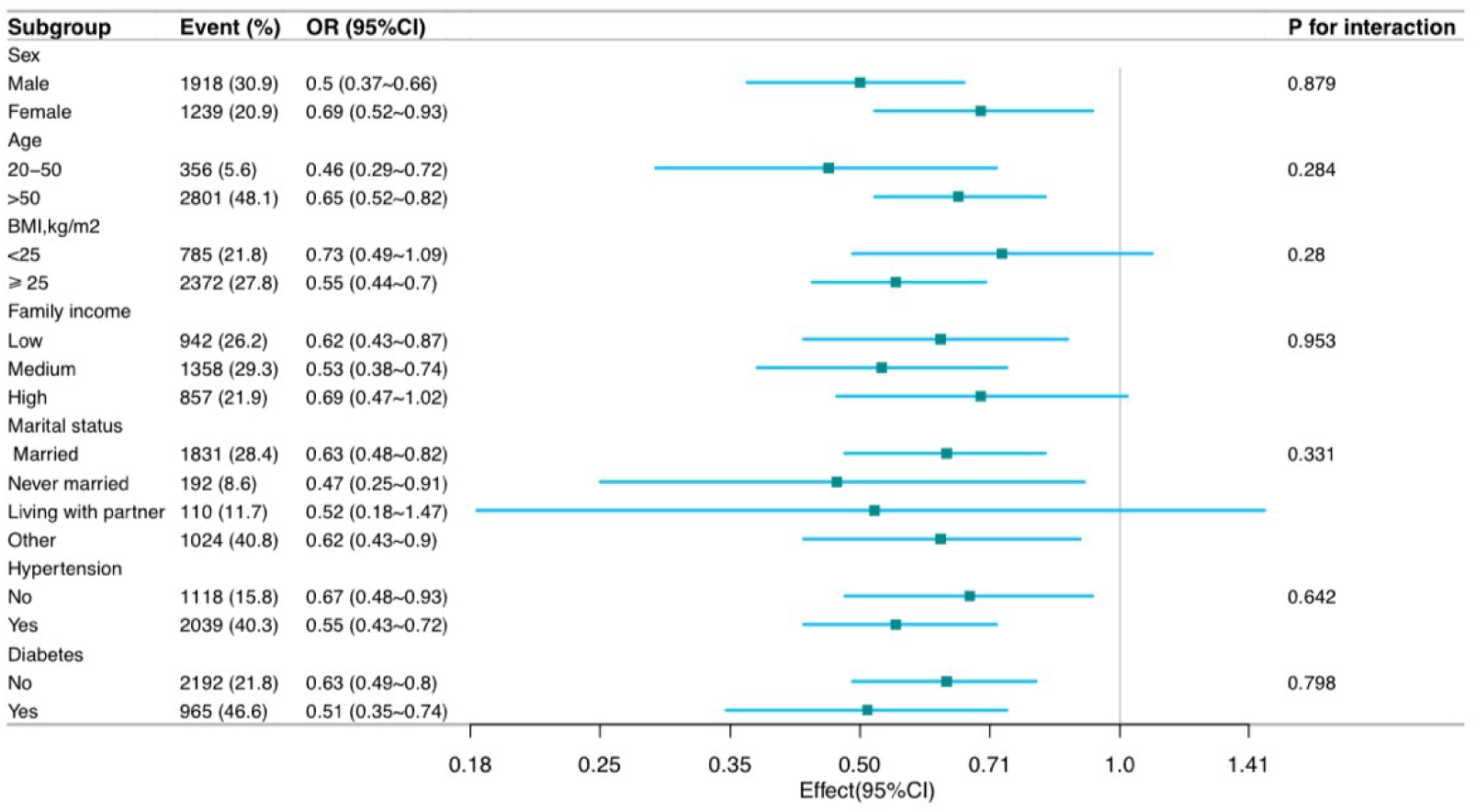
Relationship between albumin and Speech-frequency hearing loss Abbreviations:CI, confidence interval; OR, odds ratio.

**Figure 4.**
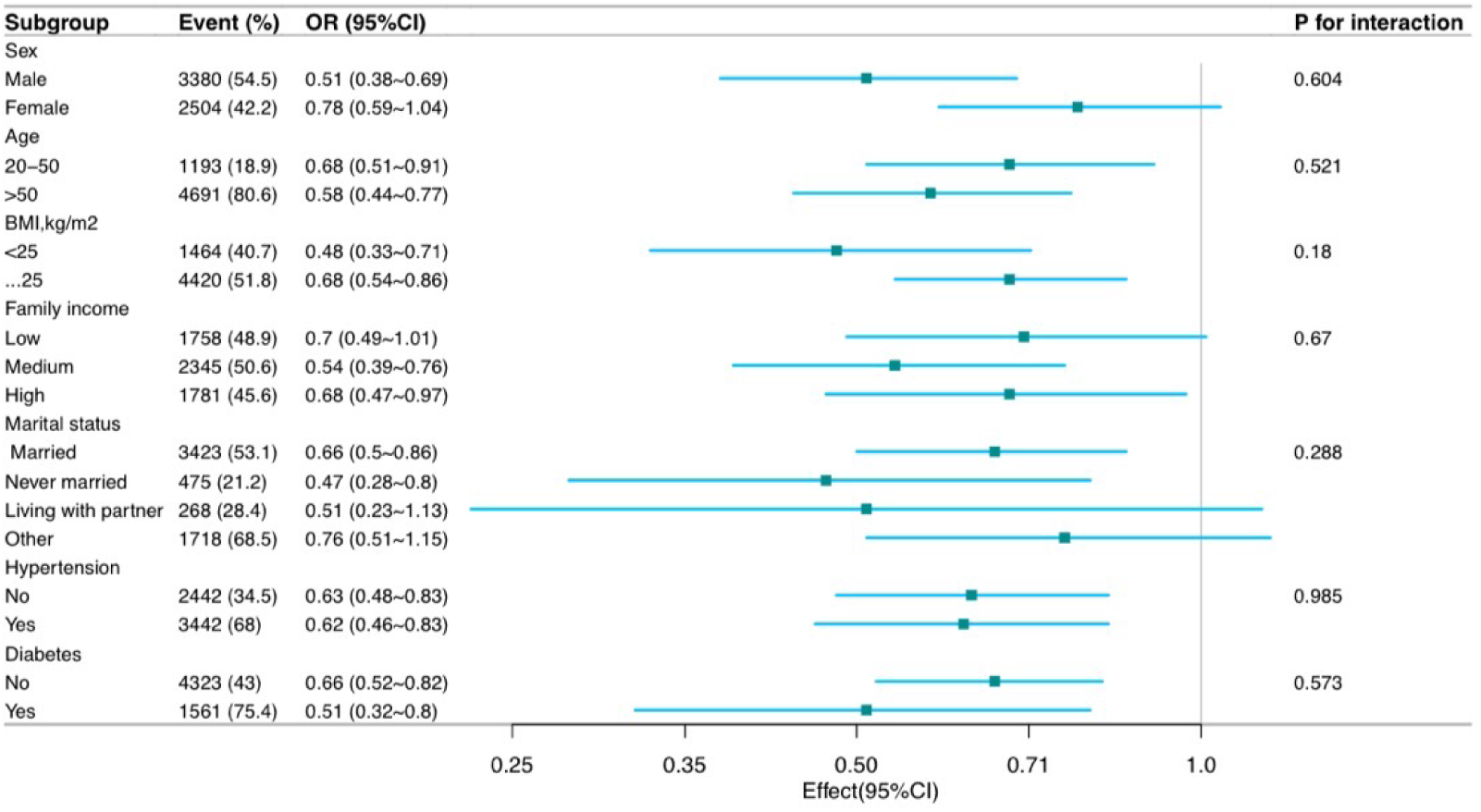
Relationship between albumin and High-frequency hearing loss Abbreviations:CI, confidence interval; OR, odds ratio.

### Sensitivity Analyses

When hearing loss exceeded 25 dB, the relationship between plasma albumin levels and hearing loss remained consistent. The adjusted ORs for plasma albumin in relation to low-frequency, speech-frequency, and high-frequency hearing loss were 0.58 (95% CI: 0.48–0.72, p < 0.001), 0.69 (95% CI: 0.55–0.86, p = 0.001), and 0.73 (95% CI: 0.57–0.93, p = 0.011), respectively (Table 4).

**Table 4.** Association between albumin and hearing loss.

| Variable | No.(Total=12133 ) | OR (95% CI) |  |  |  |
| --- | --- | --- | --- | --- | --- |
|  |  | crude | *P*-value | Adjusted | *P*-value |
| Low-frequency hearing loss( > 25dB) | 1326 (10.9%) | 0.26<br>(0.22~0.31) | <0.001 | 0.73<br>(0.57~0.93) | 0.011 |
| Speech-frequency hearing loss( > 25dB) | 2036 (16.8%) | 0.29<br>(0.25~0.33) | <0.001 | 0.69<br>(0.55~0.86) | 0.001 |
| High-frequency hearing loss( > 25dB) | 4496 (37.1%) | 0.32<br>(0.29~0.36) | <0.001 | 0.58<br>(0.48~0.72) | <0.001 |

Abbreviations:CI, confidence interval; OR, odds ratio; Ref: reference. adjusted for age, sex, race, marry, household income poverty ratio, education, smoke, alcohol, physical activity,body mass index, hypertension, diabetes, noise, glutamic-pyruvic
transaminase;glutamic oxalacetic transaminase,protein.

## Discussion

This large cross-sectional study of adults in the United States identified a substantial and consistently negative association between serum albumin levels and hearing loss across various subgroups. Notably, the findings indicate no interactive effects between albumin levels and hearing loss, suggesting consistency across subgroups, while sensitivity analyses confirmed the robustness of these results.

Zhang et al. conducted a cohort study involving 135 patients with SSNHL and 135 controls, reporting significantly lower albumin levels in patients with SSNHL. Additionally, patients with effective SSNHL exhibited significantly higher albumin levels than those with non-recovered SSNHL. These findings suggest that lower serum albumin levels are associated with the development and poor prognosis of SSNHL and that albumin may serve as a predictive marker for SSNHL[17]. A meta-analysis of 17 studies by Chen et al. demonstrated that the C-reactive protein to serum albumin ratio (CRP/Alb) was significantly elevated in the experimental group compared to controls, with a high CRP/Alb ratio indicating the occurrence of SSNHL[24].

Serkankaile et al. conducted a cross-sectional and retrospective study of 88 patients, reporting that an elevated fibrinogen-to-albumin ratio may predict poor prognosis in patients with SSNHL[25]. Similarly, Ocal et al. conducted a retrospective study of 40 patients diagnosed with idiopathic SHL and found that the C-reactivity-to-albumin ratio was significantly higher in patients with SHL than in controls, suggesting its potential utility as a prognostic indicator[26]. While previous studies have examined the relationship between albumin levels and hearing loss in relatively small cohorts, they have primarily focused on prognosis rather than investigating this association in the general population. The NHANES dataset provides a unique opportunity to explore the relationship between albumin levels and hearing loss while accounting for numerous covariates and conducting stratified analyses.

The pathogenesis of hearing loss is complex and involves multiple hypotheses, including microcirculatory disturbances[27,28], viral infection and immune impairment[29,30], oxidative stress[31–33], and endolymphatic hydrops[34,35]. While the exact cause of hearing loss remains unclear, it is likely influenced by a combination of genetic factors, noise exposure, ototoxic medications, and other environmental factors[36]. Recent research suggests that alterations in blood rheology and elevated blood viscosity may also be associated with hearing loss[37,38]. Additionally, some studies indicate that chronic inflammation may contribute to the development of hearing loss[39], as it increases the likelihood of microvascular injury and ischemia, leading to inner ear microcirculatory dysfunction[40]. Inner ear vasospasm, thrombosis, or embolism may result in ischemia, and inadequate blood supply to the cochlea can cause hypoxic-ischemic injury, mitochondrial dysfunction, oxidative stress, and cellular damage, ultimately impairing cochlear function and leading to hearing loss [41,42]. Furthermore, hearing loss may be linked to vascular disease, with affected individuals exhibiting an increased risk of cardiovascular disease[43]. For instance, high blood pressure, diabetes mellitus, and dyslipidemia have been associated with hearing loss[44]. Degenerative changes in the peripheral and central auditory nerves may also contribute to hearing impairment[45].

Albumin is a crucial blood biomarker that provides insight into physiological status and overall health. It is a sensitive indicator of homeostatic mechanisms and reflects real-time changes in the internal environment. Albumin exerts anti-inflammatory effects by inhibiting peroxidase activity and free radical production and also possesses antioxidant properties[46,47]. The cochlear vascular system is highly sensitive to inflammatory factors, and the anti-inflammatory effects of albumin may help mitigate vascular damage and preserve the blood-labyrinth barrier[24]. Oxidative stress is a key contributor to endothelial dysfunction, and vascular damage is implicated in the pathogenesis of hearing loss[31]. As a free radical scavenger, albumin binds copper and iron ions, thereby inhibiting oxidation reactions[48]. Despite these protective roles, individuals with hearing loss may experience a hypercoagulable state, leading to microcirculatory disorders in the inner ear[49]. Albumin helps maintain microvascular integrity[50], binds arachidonic acid and nitric oxide, interacts with free fatty acids, and regulates vascular tone while inhibiting platelet aggregation[13,51], thereby protecting endothelial function in the inner ear microcirculation. Additionally, albumin enhances serum osmotic pressure, attracts interstitial fluid[52], improves organ perfusion, reduces free radical production, and minimizes damage to cochlear hair cells and strial vascular cells[53]. Given these physiological properties, albumin is likely involved in the pathogenesis of hearing loss through its anti-inflammatory, antioxidant, anticoagulant, and antiplatelet aggregation activities. Its multifaceted effects on vascular endothelial function may also contribute to neuroprotection[12,54], potentially linking albumin to the neurodegenerative mechanisms underlying hearing loss. Furthermore, its roles in thrombosis, coagulation, and atherosclerosis suggest a potential connection between hearing loss and cardiovascular disease mechanisms[43,55]. Although albumin influences hearing through multiple pathways, its precise mechanisms require further investigation. Future studies integrating molecular biology approaches and clinical big data analyses are needed to elucidate the pathophysiological role of albumin in the inner ear and explore new strategies for preventing and treating hearing impairment.

Analysis of NHANES cross-sectional data provides new insights into the relationship between albumin levels and hearing loss risk. By examining this association from a unique perspective, the findings offer novelty and potential clinical relevance. The results provide strong evidence of an association between albumin levels and hearing loss risk, accounting for potential confounding factors and biases. However, several limitations should be acknowledged. First, the cross-sectional and observational design precludes definitive causal inferences regarding the interactions between albumin, covariates, and hearing loss. Additionally, despite the large sample size, the study population was limited to United States residents, necessitating caution when generalizing the findings to other populations. Future research should address these limitations to validate and expand on these findings.

## Conclusion

In adults in the United States, higher albumin levels are associated with a lower risk of hearing loss. These findings underscore the potential role of declining albumin levels as a risk factor and biomarker for hearing loss, warranting further investigation.

## Data Availability

All relevant data are publicly available from the National Health and Nutrition Examination Survey (NHANES) repository hosted by the Centers for Disease Control and Prevention (CDC). The datasets can be accessed at https://www.cdc.gov/nchs/nhanes/index.htm.

## Acknowledgements

Thanks to Dr. Liu Huanxian, Department of Neurology, First Medical Center, People’s Liberation Army General Hospital, for his contribution to this article Statistical support for manuscripts, research design consultation and comments.

